# Clinical and immunological predictors of paradoxical reactions in patients with tuberculous lymphadenitis

**DOI:** 10.1101/2025.04.01.25325023

**Authors:** A Rajendra, GJ Fletcher, JP Demosthenes, P Thangavelu, BC Ghale, R Kannangai, MS Ramesh, P Rupali

**Affiliations:** Department of Clinical Virology, Christian Medical College, Tamil Nadu, India; Department of Infectious Diseases, Christian Medical College, Tamil Nadu, India; Division of Infectious Diseases, Henry Ford Hospital Detroit, USA

## Abstract

**Background & Aim:** Tuberculous paradoxical reaction in HIV-negative patients is rare and not well understood. This study aimed to determine the incidence of paradoxical reaction and identify clinical and immunological predictors contributing to its development.

**Methods:** Patients with tuberculous lymphadenitis on anti-tuberculous therapy were monitored for paradoxical reaction. Clinical, histopathological, microbiological, and immunological parameters of paradoxical reaction (n=10) and no paradoxical reaction (n=66) groups were analysed at baseline, 2 months, and 6 months. Peripheral blood mononuclear cells from paradoxical reaction (n=9) and no paradoxical reaction (n=13) patients were stimulated with TB-ESAT peptide, and mRNA expressions of cytokines (IL-4, IL-10, IL-12, TNF-α, IFN-γ) and transcription factors (T-bet, GATA-3) were quantified.

**Results:** Paradoxical reaction incidence was 5.7% (4/70); six patients presented with paradoxical reaction at baseline. The mean paradoxical reaction onset was 3.25 months. Absence of necrosis at baseline was a significant predictor (OR: 0.069; p=0.039). IL-10 mRNA at 2 months and IL-12 mRNA at 6 months increased in the paradoxical reaction compared to no paradoxical reaction (p=0.05). In paradoxical reaction, baseline T-bet mRNA correlated with IFN-γ (r=0.79, p<0.01) and TNF-α (baseline: r=0.97, p<0.000; 2 months: r=0.86, p<0.002). In NPR, T-bet mRNA correlated with IFN-γ and TNF-α at all time points (p<0.05), while GATA-3 mRNA correlated with IL-4 and IL-10 at 2 and 6 months (p<0.05).

**Conclusion:** Differential Th1/Th2 regulation driven by transcriptional factors alters cytokine expression, influencing paradoxical reaction. Further transcriptomic/proteomic studies are needed to elucidate immune mechanisms for precise therapeutic intervention.

## Introduction

Tuberculosis caused by *Mycobacterium Tuberculosis* is the second leading cause of death from a single infectious agent (after SARS-COV-2) and is a major public health challenge(1). Tuberculous lymphadenitis (TBLN) is a common manifestation of extrapulmonary tuberculosis. Despite increasing interest and research advances, many immunological aspects of tuberculous lymphadenitis remain largely unknown(2).

Tuberculous Paradoxical Response or Reaction (PR) is an exuberant inflammatory reaction characterized by worsening of clinical or radiological findings after initiation of appropriate antitubercular therapy (ATT) in the absence of evidence of drug resistance or presence of an alternative diagnosis. This phenomenon was first recognised by Choremis et al. in children with tuberculosis who suffered a temporary worsening of fever and x-ray alterations after initiating ATT(3). Clinical recognition of this phenomenon is also fraught with difficulty as often there is worsening of underlying clinical picture i.e., suppurative/inflammatory response of involved lymph nodes or contiguous organs. A lack of response due to inadequate therapeutic drug levels of anti-tuberculosis drugs or multi-drug or extensively drug-resistant tuberculosis which may not be detected initially may also present similarly. Clues often include initial improvement and worsening clinically, lack of systemic symptoms (local features are more prominent), absence of culture positivity (lack of viable bacilli) though non-viable tuberculous bacilli may or may not be detectable by molecular techniques. Paradoxical reactions frequently mistaken as drug-resistant tuberculosis lead to needless use of toxic second-line ATT(4). Hence, predicting and diagnosing paradoxical reactions worsening is of utmost importance, as using steroids or other anti-inflammatory drugs for a short duration helps in treating the same. Most of the data regarding paradoxical worsening comes from HIV patients on highly active antiretroviral therapy (HAART)(5–7). However, limited immunopathogenic information is available for the same kind of response seen in HIV-negative patients.

Although no formal case definition has been developed, previous prospective studies have validated a consensus definition for use in clinical and research settings, which is summarised as follows(8):

A. An initial improvement after ATT initiation
B. Worsening of initial symptoms or onset of new TB-like symptoms after initiation of ATT
C. Absence of persistently active TB (due to Multidrug-resistant TB, poor compliance, impaired digestive absorption or persistently positive cultures for tuberculosis)
D. Absence of any other explanation of clinical deterioration(8)

Hence, in this prospective cohort study conducted amongst patients with tuberculous lymphadenitis, we evaluated incidence of PR, clinical predictors and correlated the same with an immunological profile to find a possible pathogenetic mechanism behind this interesting phenomenon.

## Materials and methods

This prospective study was conducted at Christian Medical College, Vellore, a tertiary care centre in India between May 2016 and June 2017. Ethical clearance was obtained from the Institutional Review Board (IRB Minute No. 9997 and 10021).

### Participants

A consecutive sampling strategy was employed for this study, wherein all patients above the age of 15 years, who presented with peripheral TBLN to outpatient and inpatient departments of Infectious Disease, Medicine and Surgery, or those already on ATT presenting within 30 days for reconfirmation of diagnosis, were also included in the study. This was considered as Cohort A or the Incident cohort. In addition, we also included patients already on ATT who presented to us with paradoxical worsening. This was categorized as Cohort B (Paradoxical reaction (PR) cohort) (Table 1, Supplementary information). Case definitions used for this trial is summarised in (Table 2, Supplementary information). A written informed consent was obtained from those eligible and willing to participate. In case of patients less than 18 years, consent was taken from the guardian after explaining the nature of the study to the patient.

**Table 1:**
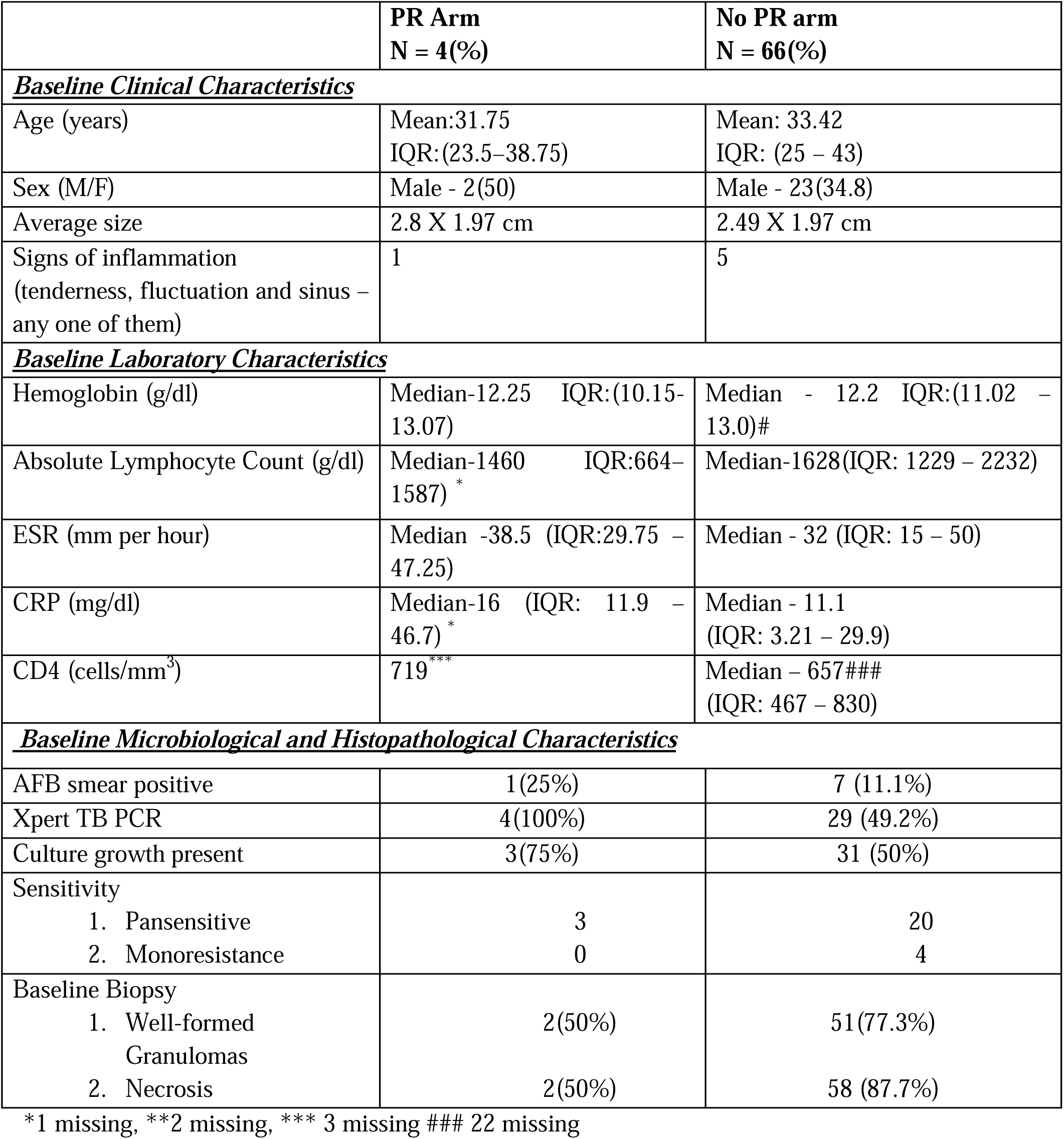
Comparison between PR arm and Non PR arm in the incident cohort.

**Table 2:**
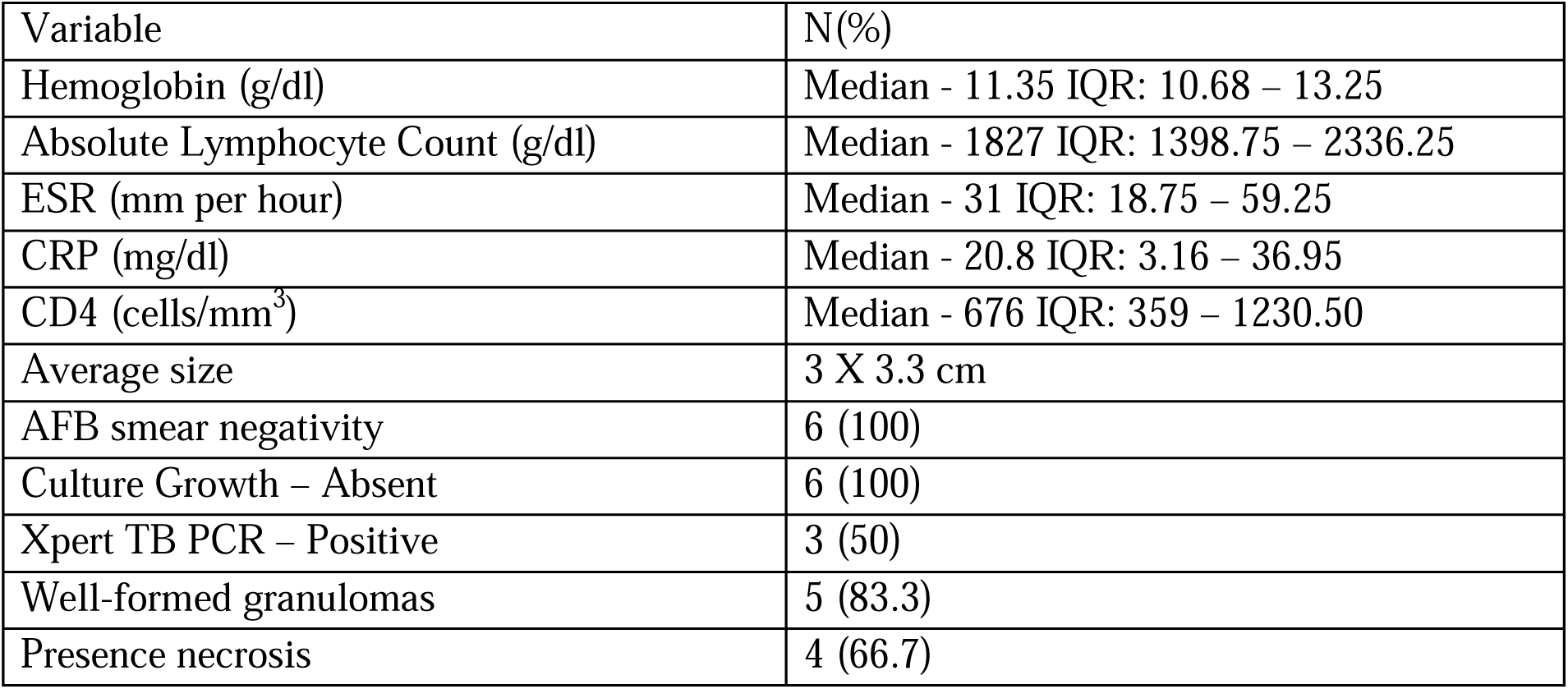
Baseline Characteristics of Patients presenting with Paradoxical(PR group) Worsening.

### Clinical assessment

At initial presentation, baseline lymph nodal groups were examined to note the site, size, number of involved lymph nodes along with presence of signs of inflammation (local tenderness, fluctuation and discharge). Photograph of the involved lymph node was taken for future comparisons. Baseline laboratory parameters which were assessed included haemoglobin, erythrocyte sedimentation rate (ESR), C - reactive protein (CRP), white blood cell (WBC) count, differential Count, absolute Lymphocyte count (ALC) and CD4 count.

The study participants were reviewed at the end of intensive phase (2 months) of ATT and then at end of treatment. At each follow-up visit, the involved lymph nodal areas were examined, and other lymph nodal areas were examined to look for development of new nodes. Clinical parameters like lymph node size, site, number and presence/absence of signs of inflammation were recorded. Laboratory parameters were repeated at 2 months and end of therapy.

At the time of diagnosis of PR, poor drug compliance and drug resistance were evaluated. We performed a pill count, followed up previous cultures to rule out resistance and if there was a doubt of the same, repeat biopsy was advised for confirmation of the diagnosis and performed if the patient consented. In case of lymph node abscess if surgical aspiration/debridement/drainage was planned for therapeutic purposes, sample was sent for mycobacterial PCR and/or culture positivity.

### Immune Profiling of Patients

Ten ml of blood was collected for all the patients at baseline, 2 months and 6 months as well as at the time of PR for evaluation of cytokine profiles. CD4 + T cell enumeration was done using a flow cytometer (BD FACS Count, CD4 reagents, San Jose, USA) on all samples at baseline. Estimation of inflammatory markers, namely C-reactive protein (CRP) was performed with the siemens nephelometry assays atellica NEPH 630 System (Erlangen, Germany). The following cytokines and transcription factors were measured: Cytokines-IL-4, IL-10, IL-12, IFN-γ and Transcription factors-T-bet, GATA-3. Peripheral Blood Mononuclear Cells (PBMC) were separated using the isopaque-ficoll gradient centrifugation method. PBMC were stimulated with early secreted antigenic target 6 kDa (ESAT) (5 µg/ml), for 48 hours or medium alone as a negative control at 37°C with 5% CO2. The cells were harvested and stored at -70°C. Subsequently the cells were subjected to mRNA extraction and the quantification of mRNA of the cytokines and relevant transcription factors (T-bet/GATA-3) were done using PCR Array (Qiagen Sciences, Maryland USA). The PCR was performed in AB 7500 real time PCR (Applied Biosystem, USA). PCR cycling conditions were set as follows: 95 °C for 10 min, 40 cycles of 95 °C for 15 s and 60 °C for 30 s. The relative changes in gene expression were calculated using comparable threshold cycles - ΔΔCt method. Data was analysed by a web-based analysis program RT2 profiler PCR array data analysis version 3.5 from SA Biosciences. We hoped to compare clinical, laboratory and immunological profiling of PR patients and compare it with those with NPR.

### Endpoints

The primary endpoint was to determine the proportion of patients developing PR in the incident cohort. Secondary endpoints were to identify clinical, laboratory parameters and immunological parameters which could be associated with or predict PR.

### Sample Size

With a presumption that the incidence of PR is 10-15% the required sample size was 138. However, in view of slow accrual the study was stopped after 80 patients.

### Statistical Analysis

Data entry was done using EPIINFO Software. PR group included patients from the incident cohort who developed PR while on follow-up and patients from the PR cohort. Frequencies of clinical and laboratory parameters between patients who developed PR (PR group) were compared with those who did not develop PR (No PR group). A univariate analysis was done using a chi-square test for dichotomous variables and a students’ T test for continuous variables. For the cytokine profile, pearson’s correlation was conducted to find the relationship between the transcription factors and the signature cytokines. All statistical analyses were performed using the medcalc software version 9.2.0.1, graph pad prism (version 6.0.1). Statistical significance was set as *P* < .05. Additionally, RStudio was used to generate the correlation heatmaps.

## RESULTS

Between May 2016 through June 2017, 80 patients, who fulfilled the inclusion criteria were screened. Eventually, 76 patients were included in the final analysis, the reasons are detailed in the flow chart (Figure 1). Seventy patients belonged to the incident cohort and 6 patients presented with PR (PR cohort).

**Figure 1:**
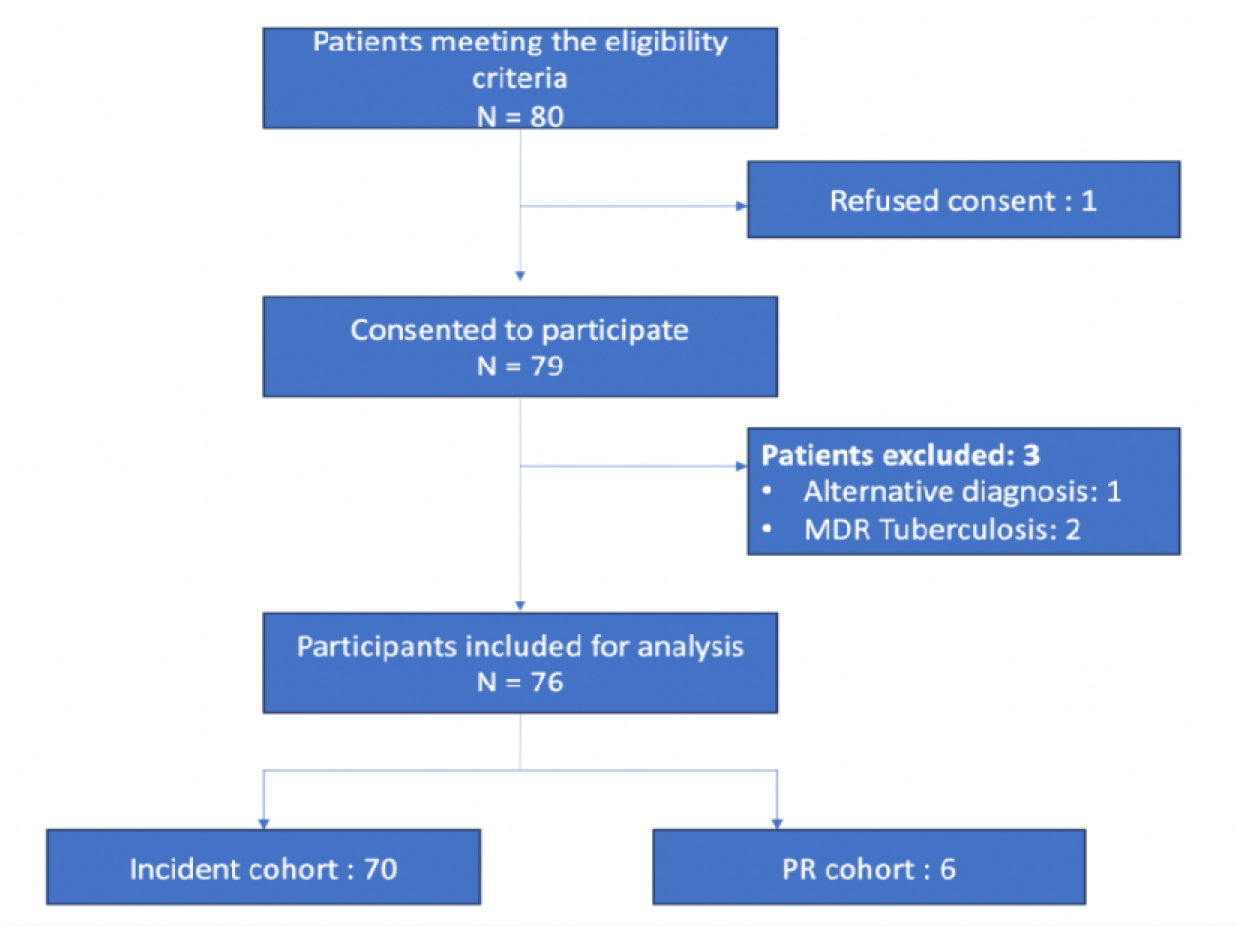
STROBE diagram summarising the recruitment of patients.

### Incident cohort

#### Baseline characteristics of the incident cohort (N = 70)

Of the 70 patients, 25 were men and 45 were women. Mean age of the cohort was 33.3 (IQR: 25 – 42.25) years. About 10% of patients were on ATT at the time of recruitment. Exclusive peripheral lymph nodal enlargement was seen in 72% and 28% had additional organ involvement. Baseline demographic, clinical, laboratory, microbiological and histopathological characteristics of these patients is summarized in Table 3, Supplementary information. Only 8 patients (11.4%) had a positive AFB smear from the lymph node biopsy. Xpert TB PCR was positive in 33 patients (47.14%) and none of them had rifampicin resistance. Mycobacterial culture was positive in 34 patients (48.57%). Amongst the patients in whom sensitivity results were available, 23 were pan-sensitive. 4 patients had resistance to at least 1 drug, however, none of them had multidrug-resistant or extensively drug-resistant *M. tuberculosis*. Lymph node histopathology showed well-formed granulomas in 53 patients (75.71%) and necrosis in 60 patients (85.71%). Most involved lymph nodes were cervical nodes. In most instances, there was 1 lymph node group involved. 8 patients had tenderness and fluctuation, 1 had discharge and 5 had sinuses at baseline.

**Table 3:**
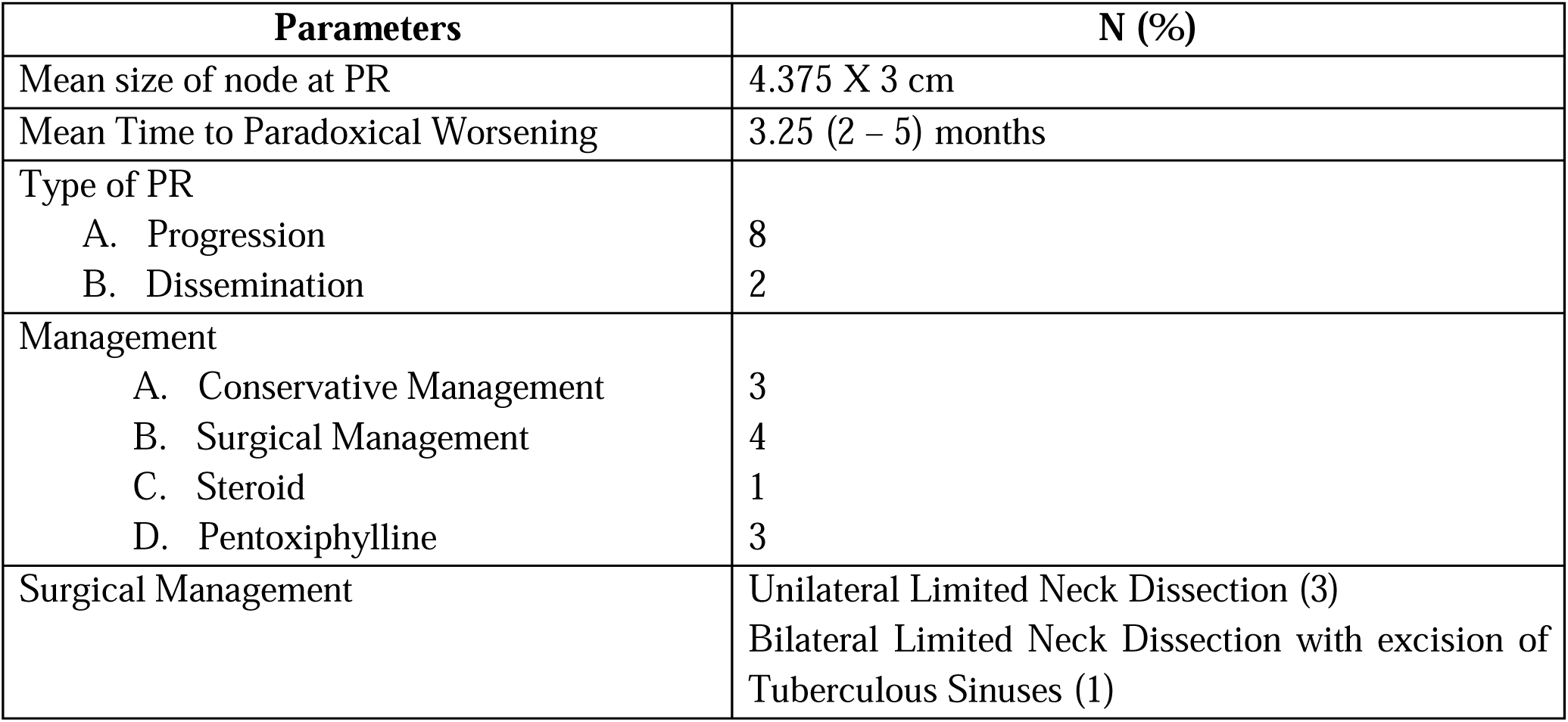
Treatment of patients with PR in both cohorts.

### Comparison between the patients who developed and did not develop PR in the incident **cohort**

In the incident cohort (n = 70), 4 patients developed PR. The incidence of PR was 5.7%. Clinical, laboratory, microbiological and histopathological features of the patients who developed PR (4 patients) were compared against those who did not (66 patients) (Table 1). Among the PR patients, only 1 (25%) had signs of inflammation at baseline compared to 5 (7.6%) in the No PR group. At baseline, mean size of lymph node, hemoglobin, absolute lymphocyte count, ESR and CRP were similar in both the groups. CD4 count was available in 1 patient with PR which was 719 cells/mm^3^. Median CD4 count for the No PR patients was 657 cells/mm^3^(available in 44 patients). There was no significant change in the median values of CRP when measured at the end of intensive phase, at the time of PR and at the end of continuation phase between the two groups (Figure 2).

**Figure 2:**
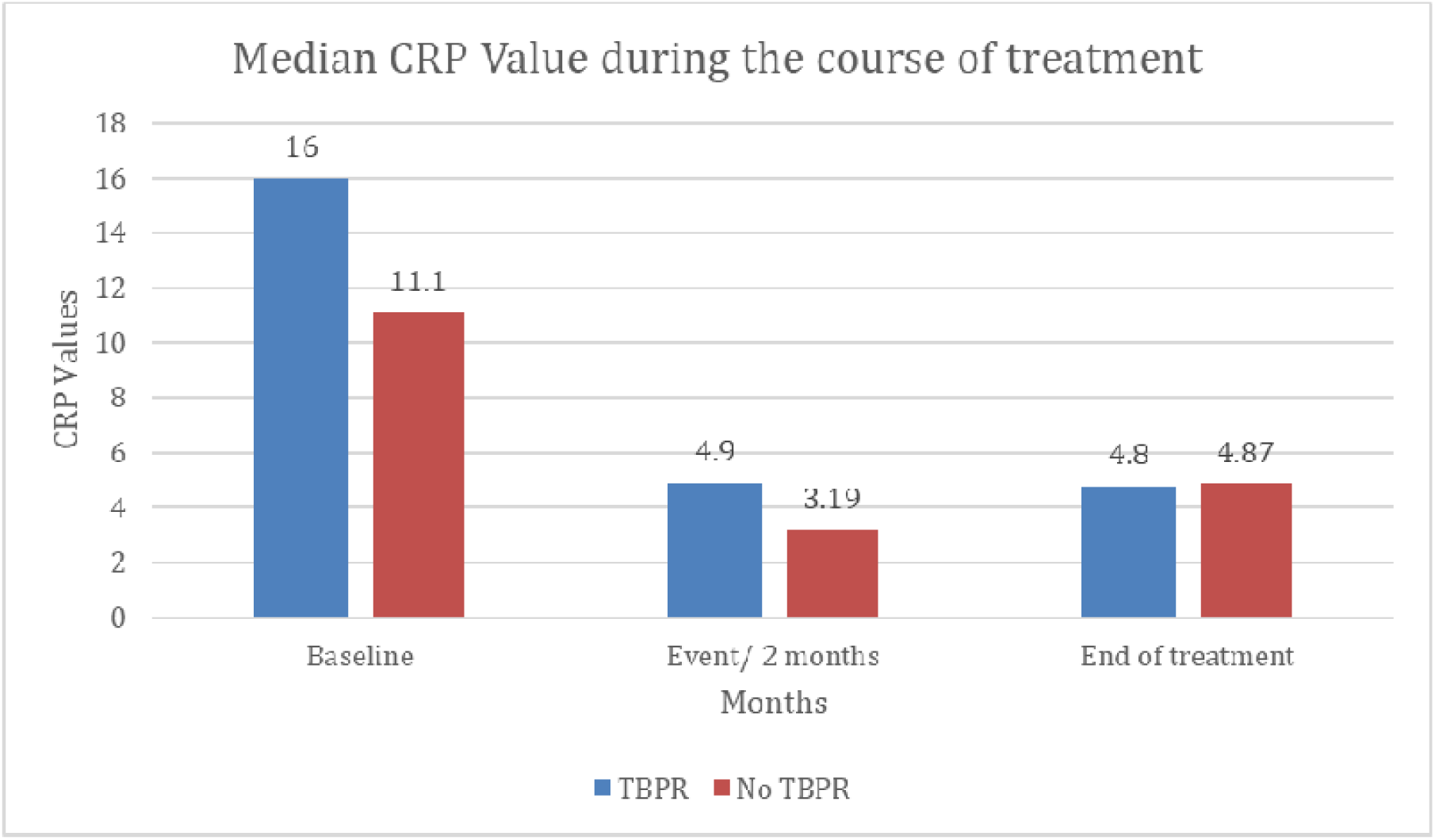
Comparison of CRP levels among the Paradoxical and No-Paradoxical group at thr e different time points (Baseline, 2m and 6m of treatment)

### Univariate analysis of factors associated with development of PR in the incident cohort

By univariate analysis, the only significant factor was presence of necrosis. Presence of necrosis negatively predicted the occurrence of PR (Odds ratio: 0.069(95%CI: 0.008 – 0.626; P 0.039). None of the other clinical, laboratory, microbiological or histopathological factors could predict the occurrence of PR. Univariate analysis is summarised in (Table 4, supplementary information).

### PR Cohort (n = 6)

6 patients were in the PR cohort, who satisfied the diagnostic criteria for PR at presentation itself. Half of these patients had signs of inflammation, and the rest presented with an increase in the node size from before (based on patient history). The average size of the nodes at presentation was 3 X 3.3 cm. The baseline parameters of the PR cohort are summarised in (Table 2).

### Treatment of those with PR in both cohorts

Overall, 10 patients were diagnosed with paradoxical worsening, 4 of them in the incident and 6 in the PR cohort. The mean time to paradoxical worsening was 3.25 months [Range: 2 – 5 months]. One patient was managed with steroids, and 3 with pentoxifylline. Four patients required surgical management, which included neck dissection and excision of necrotic lymph nodes and sinuses. Three patients were managed conservatively by continuing ATT. Treatment characteristics are summarised in (Table 3).

### Immune Profile Assessment

Comparable CD4+ T-cell counts were observed between patients with PR and those without PR. Immune cytokine profile was available in 9 patients with paradoxical reaction (PR) and 13 patients with no paradoxical reaction (NPR). Comparative analysis of immune response between the two groups revealed no significant differences observed in baseline mRNA expression of immune markers like cytokines (IFN-γ, IL-4, IL-10, IL-12 &TNF-α) and transcription factors (T-bet, GATA-3) indicating stable cytokine modulation. The PR group had minimal immunomodulatory changes (<2-fold), while the No PR group showed significant upregulation (≥2-fold) in all immune markers’ mRNA expression during treatment.(Figure 3a – 3b, and supplementary figures 3(3.1-3.6)in the PR cohort vs S1-7b in “no PR” cohort).

**Figure 3a:**
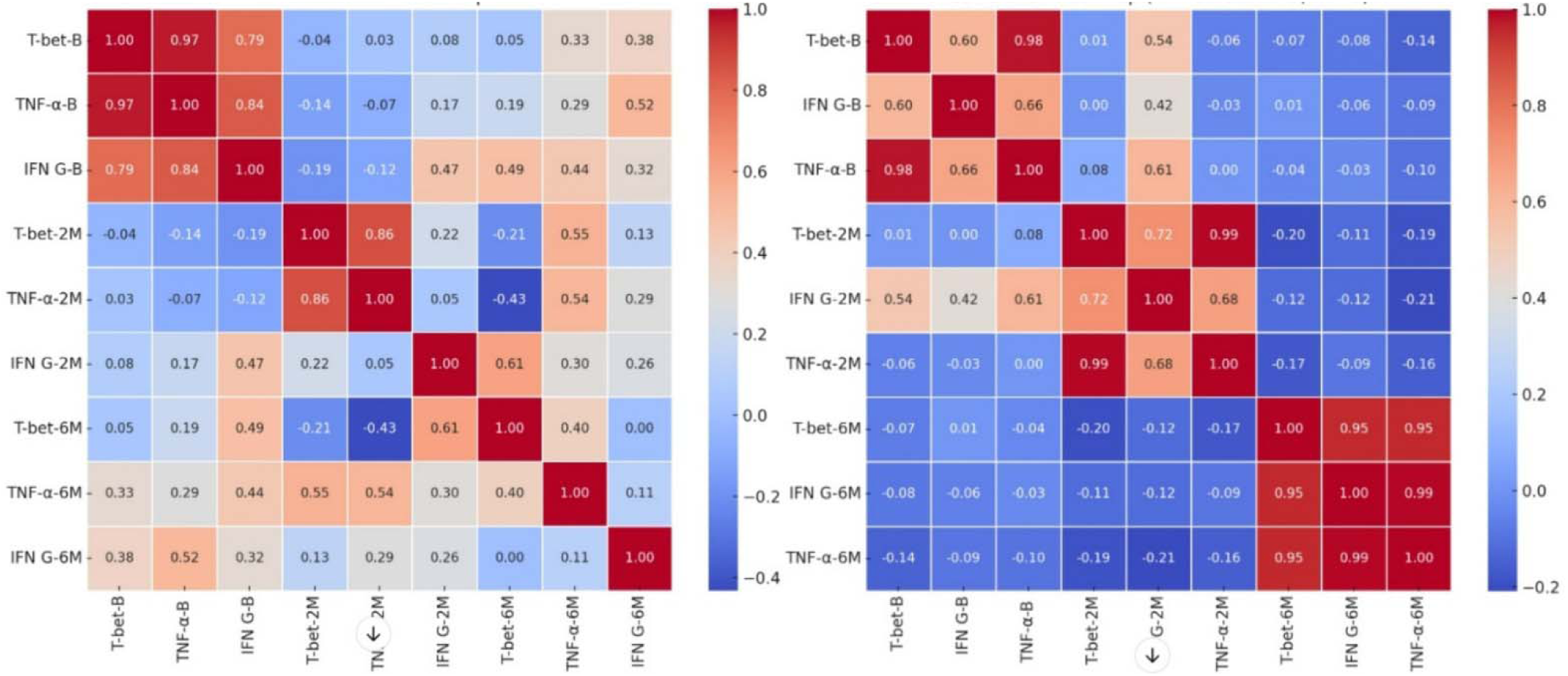
Differences in correlation between T-bet and Th1 cytokines (IFN-γ and TNF-α) in paradoxical and No-paradoxical reaction groups. Correlation of Transcription factor **T-bet** with signature cytokine of Th1 immune response IFN-γ and TNF-α in the Paradoxical Group (PR-9) vs Non-Paradoxical Group (NPR-13) PR: Paradoxical Group (n-9), NPR: Non-Paradoxical Group (n-13), Correlation coefficient – r,

**Figure 3b:**
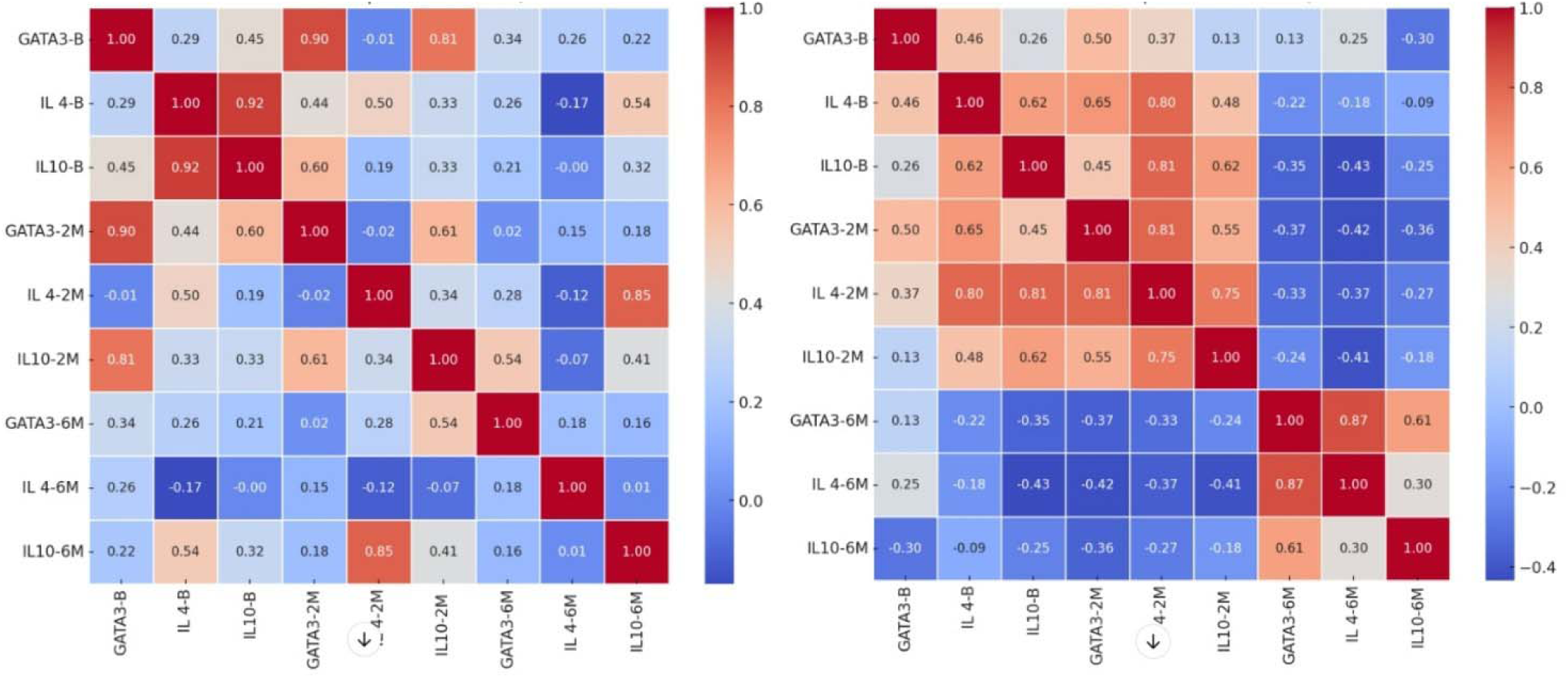
Differences in correlation between GATA-3 and Th2 cytokines (IL-4 and IL-10) in paradoxical and No-paradoxical reaction groups. Correlation of Transcription factor **GATA-3** with signature cytokine of Th2 immune response **IL-4** and **IL-10** in the Paradoxical Group (PR-9) Vs No-Paradoxical Group (PR-13) PR: Paradoxical Group (n-9), NPR: Non-Paradoxical Group (n-13), Correlation coefficient – r.

Correlation analysis uncovered strong positive associations indicating coordinated transcriptional regulation. We tried to correlate transcription factors with the signature cytokine of Th 1 and Th 2-immune responses in those with PR vs those with No PR (Figures 3a and 3b) (Tables 5 and 6, supplementary information). We found no correlation between transcriptional factor T-bet with IFN-γ in the PR group, however there was a significant correlation between T-bet and IFN-γ at all time points in the No PR. In addition, there was a significant correlation between T-bet and TNF-α at at all time points in the No PR group and in the PR group this correlation was significant only at baseline and 2 months. Overall, there seemed to signify a preferential correlation of T-bet with TNF-α rather than IFN-γ. Similarly, transcription factor GATA 3 and signature cytokines of Th 2 immune responses IL-4 and IL-10 both showed a significant correlation with GATA-3 in the No PR group, but this was not seen in the PR group suggesting that there was a differential regulation of cytokine expression influencing the polarisation of T-cell response in PR vs No PR group. A radar map compared mean cytokine levels of PR and No PR groups over time, illustrating immune response patterns. (Figure 2 Supplementary Information)

## Discussion

In this prospective study of patients with TBLN, the incidence of PR was 5.7%. Previous reports on the incidence of PR in non-HIV patients range from 2.3% to 23% (4,8–12). Patients with PR in our study presented with enlarged lymph nodes, newer nodes accompanied by development of signs of inflammation. The most common presentation was that of progression at the primary site rather than at distant sites.

PR is an exuberant inflammatory response seen after initiation of antitubercular therapy, similar to immune reconstitution syndrome (IRIS) seen in HIV-positive patients(7). The underlying pathogenesis is not fully understood. At the time of TB infection, the immune response is typically a T-helper-2(Th-2) response which is anti-inflammatory. As treatment progresses the immune response transitions to a T-helper-1(Th-1) response which is pro-inflammatory. An overwhelming immune-restitution (Th-2 to Th-1) results in the development of paradoxical worsening(13,14). In our cohort, amongst 10 patients who developed PR, AFB smear and culture results were negative in all but one patient. Histopathology at the time of PR revealed well-formed granulomas (87.5%) with necrosis (75%) in most patients, highlighting the exaggerated host immune response. Over half of these patients had Xpert MTB/RIF positivity at diagnosis of PR, possibly due to the detection of non-viable bacilli.

Several risk factors have been suggested for PR in literature which includes: younger age at presentation(10,12), male sex(10,12), lymph node tuberculosis(15), lymph node size >3 cm(11), tenderness at the time of diagnosis(10), associated extra-lymph nodal tuberculosis(4,11,16), lymphopenia at baseline(17), greater change in lymphocyte count(17), elevated baseline monocyte count(12), anaemia(17), hypoalbuminemia(17) and positive TB culture(15). However, in our study, we did not find any significant association between these risk factors and development of PR. Interestingly, we found presence of necrosis on histopathology of lymph nodes at baseline to be associated with a lower likelihood of development of PR. This suggests that patients with necrosis at baseline have a more balanced Th1/Th2 response, reducing the risk of drastic immune shift leading to PR. One of the major limitations in our study is the small number of PR events, which would have limited our ability to identify significant associations.

Among those who developed PR, immunomodulation with steroids(n=1) and pentoxiphylline (n=3) was done in 4 and in others continuation of ATT was sufficient. Four patients required surgical debridement with neck dissection and excision of necrotic tuberculous lymph nodes/ sinuses. Patients managed surgically responded better and did not require further immunomodulation. Surgical management seemed to eliminate the local inflammatory arc more effectively reducing the need for immunomodulation, reducing symptoms/signs as well as prolongation of anti-tubercular therapy. This has been demonstrated in a previous PR series published by our institution(18).

Overall, our preliminary findings indicate there is a definite signal towards a differential expression of cytokines and transcriptional factors of both Th 1 and Th 2 immune responses in the group which had a PR vs the group which did not i.e., No PR. However, there were no definitive associations between the transcription factors *t-bet* and *GATA-3* (which regulate Th-1 and Th-2 cytokine response) or cytokines (IL-4, IL-10, IL-12 and IFN-γ) with the occurrence of PR. T-bet correlated strongly with IFN-γ and TNF-α at baseline and 2 months, but this was lost at 6 months suggesting that there was a mitigation of the inflammatory response either due to the natural history or intervention due to some anti-inflammatory agents. In the No PR group, we noted a strong correlation with T-bet and TNF-α and IFN-γ at all time points suggesting a protective Th-1 immune response. On immune profiling of signature markers for Th-2 response, there was a significant correlation between GATA-3 and IL-4 and IL-10 in the NPR group at 2 and 6 months, but this was not observed in the PR group. However, a more comprehensive cytokine profiling is needed to validate this phenomenon.

The significant correlation of mRNA levels of T-bet and IFN-γ seems similar to the MAC-IRIS mouse model data reaffirming that CD4+ T-cells and IFN-γ production can influence disease, leading to the expansion of Mtb-specific Th1 cells even in non-HIV paradoxical reactions in active TB patients(13,19).

A strong correlation of T-bet with the mRNA levels of TNF-α at baseline and two months was also observed supporting evidence that the right amount of TNF-α is protective i.e., too little or too much production can drive disease as seen in *Mycobacterium marinum* infection of zebra fish (*Danio rerio*), which may enhance an appreciation of PR/IRIS(19,20) or what is known as the ’Goldilocks effect’(21,22). Following the effect of the increased or decreased production of TNF-α, there is necrosis of macrophages and un-inhibited extracellular bacterial replication. The potential relevance of this phenotype to human TB is confirmed by genetic studies of homologues of implicated factors(22). Our study findings suggest that T-bet and GATA-3-driven TH-1/TH-2 polarisation and the expression of respective TH1/TH2 cytokines as a result may be associated with the development of paradoxical reactions.

### Limitations

This was a preliminary pilot study done on a limited number of patients and will need to be studied in a larger cohort of patients. It is also likely that measurement of these cytokines in tissue rather than in serum may be a better indicator of an aberrant immune response. However, detailed transcriptomic and proteomic studies are warranted to elucidate the underlying pro-inflammatory immune mechanisms towards precise therapeutic intervention.

## Data Availability

All data produced in the present study are available upon reasonable request to the authors

## Acknowledgement

We acknowledge the invaluable support and contributions of the Department of Medicine, the Department of Clinical Microbiology, and the Department of Surgery. Their guidance, resources, and collaboration were instrumental in completing this study.

---

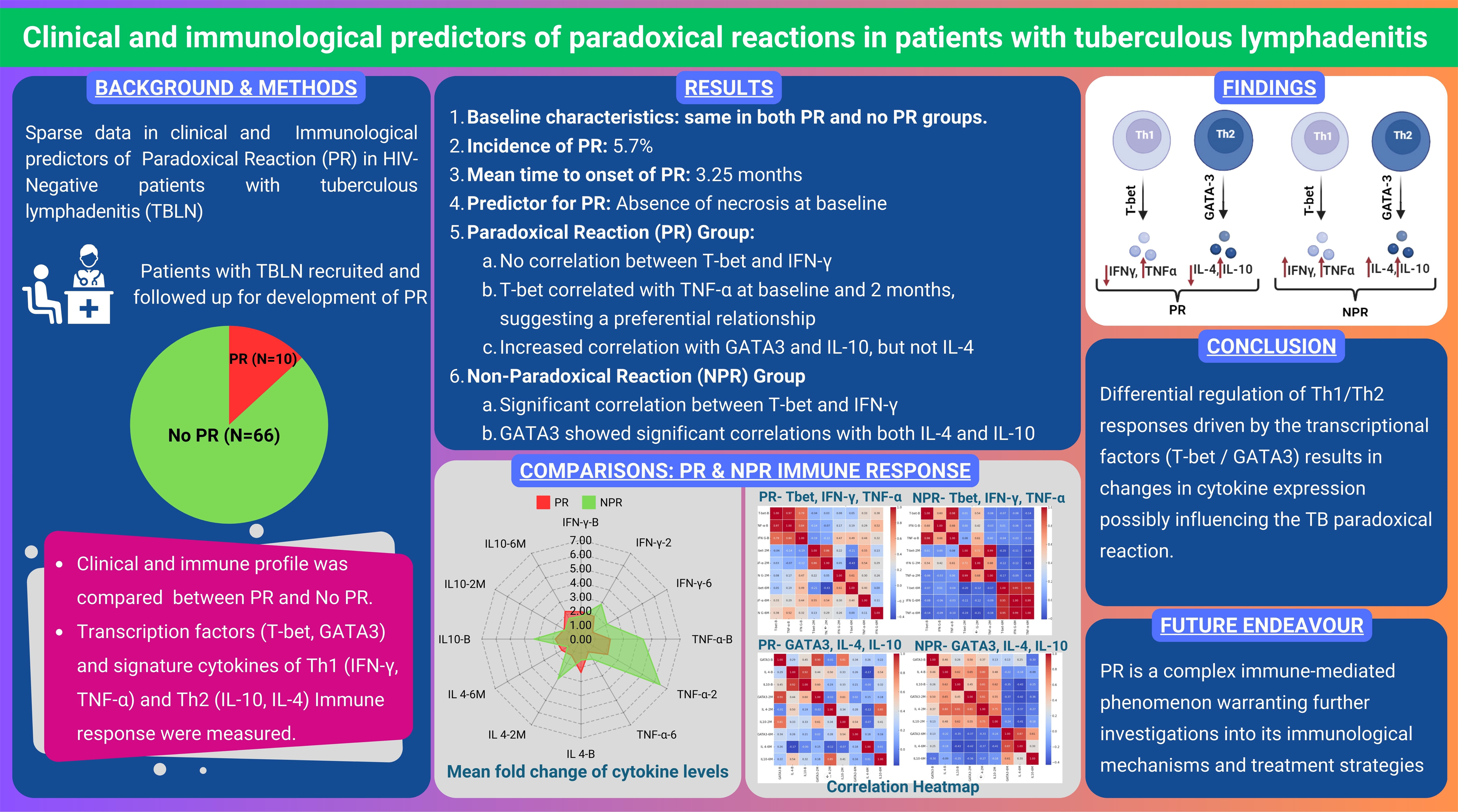

## Notes

### Competing Interest Statement

The authors have declared no competing interest.

### Funding Statement

This study was funded by CMC Internal Fluid Grant

### Author Declarations

Ethics committee/IRB of Christian Medical College Vellore gave ethical approval for this work

