## Supplementary Figures for "Clinical and immunological predictors of paradoxical reactions in patients with tuberculous lymphadenitis"

**FIGURE 1: Comparison of m RNA levels of cytokines(IL-4, IL-10, IL-12, TNF-α, IFN-γ) among the paraxodial and non-paradoxial groups at three different time points(Baseline,2m and 6m of treatment)(Supplementary Information)**

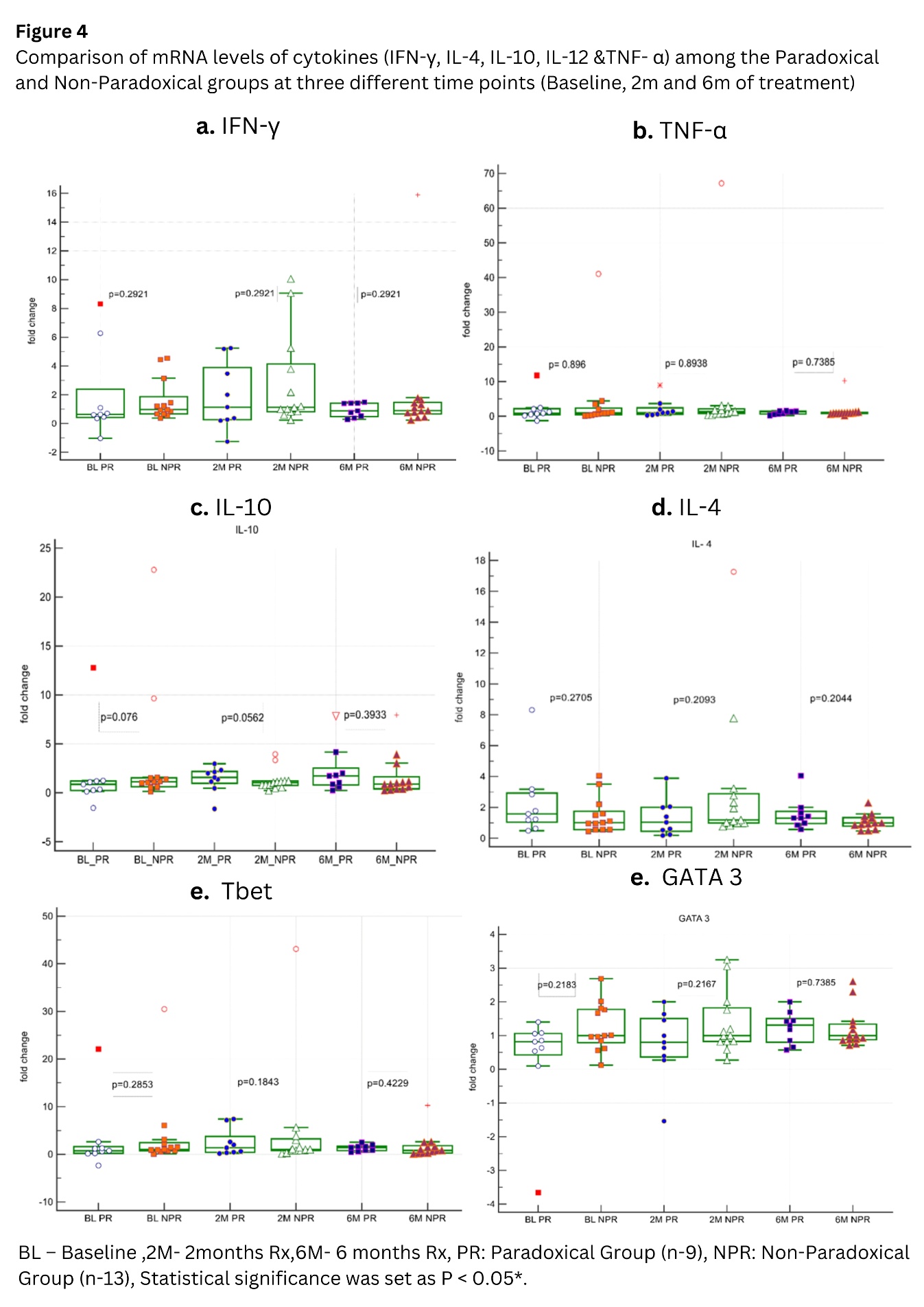

**FIGURE 2: Comparison of Mean cytokine levels between No PR and PR (Supplementary Information)**

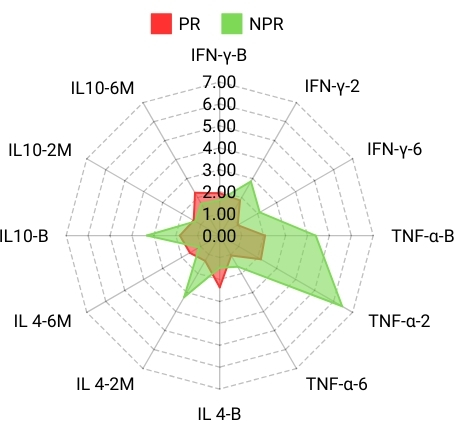

**Fold change of signature cytokine in the paradoxical reaction cohort (n=9) and**

**in the no paradoxical reaction cohort (n=13) (Supplementary Information)**

**Figure 3.1: Level of INF-γ in those with PR vs in those without PR**

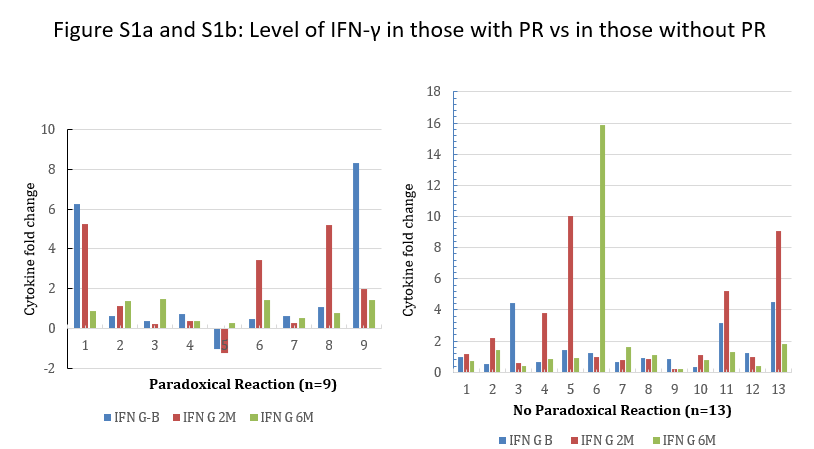

**Figure 3.2: Level of IL-12A in those with PR vs in those without PR**

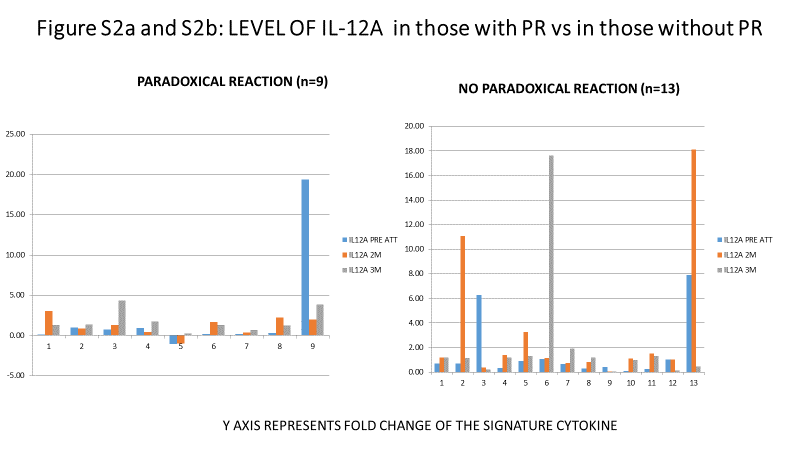

**Figure 3.3: Level of TBX21 in those with PR vs in those without PR**

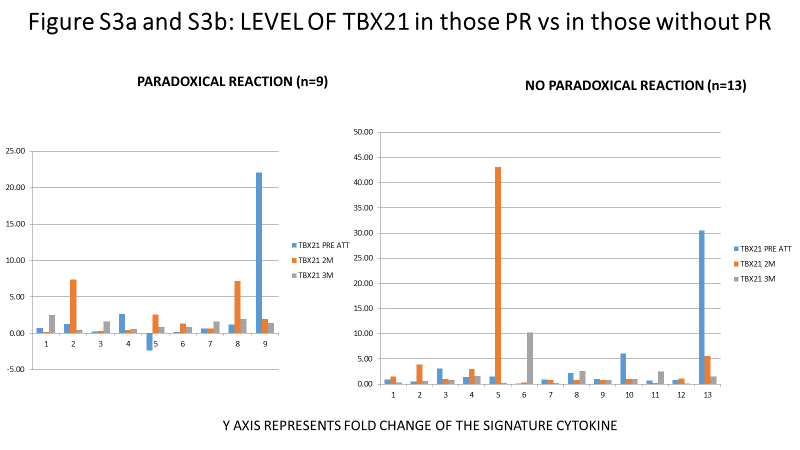

**Figure 3.4:Level of TNF- α in those with PR vs in those without PR**

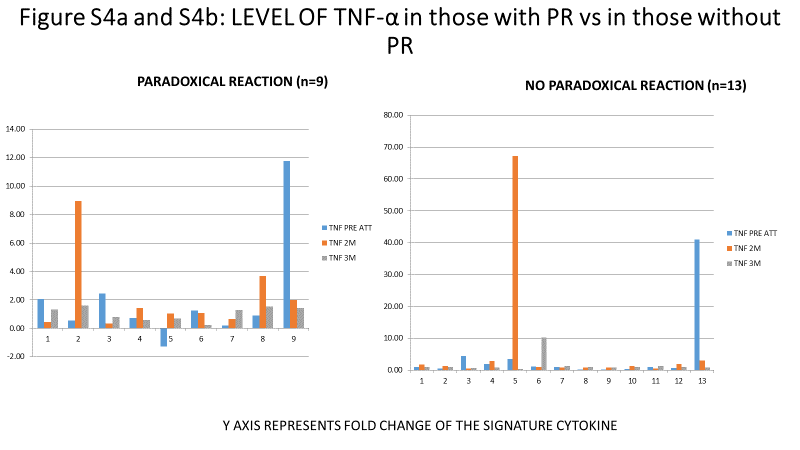

**Figure 3.4: Level of IL-4 in those with PR vs in those without PR**

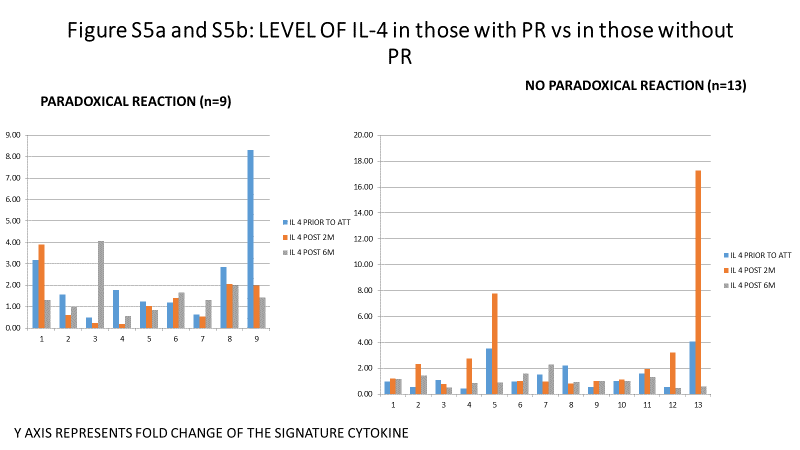

**Figure 3.5: Level of IL-10 in those with PR vs in those without PR**

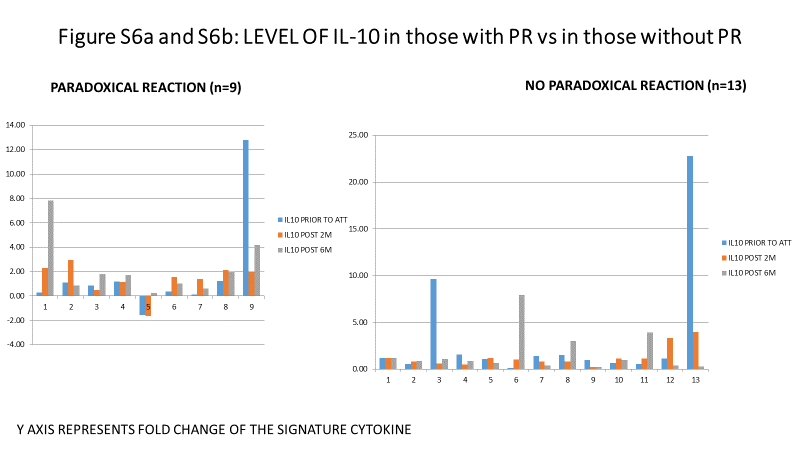

**Figure 3.6: Level of GATA-3 in those with PR vs in those without PR**

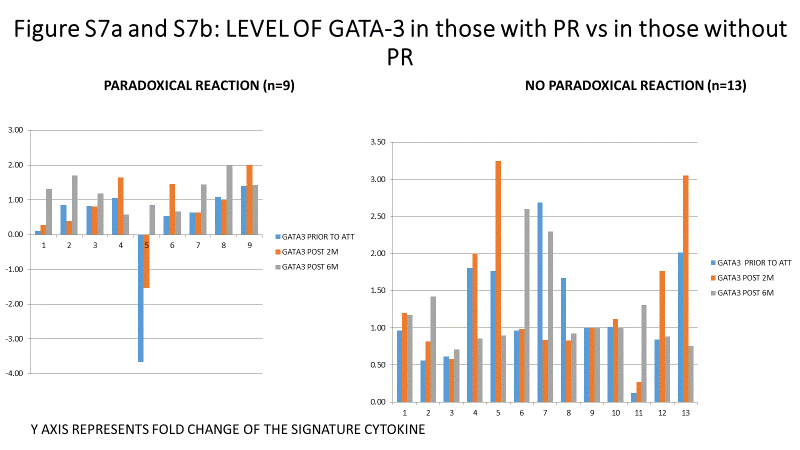
