## Supplementary Tables for "Clinical and immunological predictors of paradoxical reactions in patients with tuberculous lymphadenitis"

**Table 1: Inclusion and exclusion criteria (supplementary information)**

**Inclusion criteria**

Patients > 15 years with peripheral TBLN

Planned to be initiated on ATT

Patients who were already on ATT for less than 30 days were also included

Patients presenting with paradoxical worsening

**Exclusion criteria**

Isolated Mediastinal or abdominal lymph nodal involvement with tuberculosis

Human Immunodeficiency Virus (HIV) co-infection

Multidrug resistant tuberculosis (MDR) or extensively drug-resistant tuberculosis (XDR) as identified on tissue sample by molecular or culture techniques

Patients with connective tissue disorder or malignancies or immunodeficiency

Patients who have been on steroids and immunosuppressants

**Table 2: Case definitions (supplementary information)**

***Confirmed Tuberculous Lymphadenitis(TBLN):***

Patients were considered as confirmed TBLN when the patient had characteristic clinical findings, histopathological findings of tuberculous lymphadenitis with polymerase chain reaction(PCR)/culture positivity for Mycobacterium tuberculosis in the tissue sample.

***Probable Tuberculous Lymphadenitis:***

Patients were considered as probable tuberculous Lymphadenitis when clinical and histopathological findings were characteristic of tuberculous lymphadenitis with negative microbiology and positive response to therapy.

***Tuberculous Paradoxical Reaction/ Worsening (TBPR):***

Patients with tuberculous lymphadenitis were diagnosed to have TBPR if the patient fulfilled the following diagnostic criteria mentioned above(8).

### Table 3: Baseline characteristics of the incident cohort (N = 70) (Supplementary information)

| ***Demographic Parameters*** | ***N(%)*** |
| --- | --- |
| Sex | Male – 25 (35.7); Female – 45 (64.3) |
| Age (years) | Mean - 33.33; IQR: 25 – 42.25 |
| Patients previously treated with ATT | 7 (10) |
| Extra-lymph nodal involvement | 19 (27.1) |
| Patients who were on prior ATT | 7 (10%) |
| ***Clinical characteristics*** | ***N(%)*** |
| Site of Lymphadenopathy | Right Cervical: 48  Left Cervical: 35  Right Axillary: 4  Left Axillary: 1  Inguinal: None |
| Lymph node size | Mean - 3.594 X 2.807 cm |
| Signs of inflammation at presentation | Yes - 14(20)   - Tenderness: 8 - Fluctuation: 8 - Discharge: 1 - Sinus: 5 |
| ***Laboratory Parameters (n=45)*** | ***N(%)*** |
| Hemoglobin (g/dl) | Median: 12.22; IQR: 11.03 – 12.20 |
| Absolute Lymphocyte Count (g/dl) | Median: 1587 ; IQR: 1216 – 2231 |
| ESR (mm per hour) | Median: 36; IQR: 16 -50 |
| CRP (mg/dl) | 11.9; IQR: 3.35 – 30.9 |
| CD4 (cells/mm^3^) | Median: 662; IQR: 476 – 822 |
| ***Microbiological Parameters*** | ***N(%)*** |
| AFB smear positive | 8 (11.4%) |
| Culture Growth – Present | 34(48.6%) |
| Xpert TB PCR – Positive | 33 (47.1%) |
| Pansensitive organism | 23 |
| Resistance to one drug (H or S) | 4 |
| ***Histopathological characteristics*** | ***N(%)*** |
| Well-formed granulomas | 53 (75.7%) |
| Presence of necrosis | 60 (85.7%) |

**ATT: Anti-tubercular therapy, IQR: Interquartile range, H =Isoniazid, S = Streptomycin**

**Table 4 - Univariate analysis in the incident cohort (supplementary information)**

| **Variable** | **PR**  **N = 4**  **(Median)** | **Non – PR**  **N = 66**  **(Median)** | **Odds Ratio** | **95% CI** | **P value** |
| --- | --- | --- | --- | --- | --- |
| ***Clinical Variables at baseline*** | | | | | |
| Age < 40 | 4 | 46 | 0.920 | 0.848 – 0.998 | 0.319 |
| Sex (M/F) | 2/2 | 23/43 | 1.87 | 0.27 – 14.154 | 0.613 |
| Size >2 cm at baseline | 1 | 7 | 0.57 | 0.05 – 6.27 | 0.530 |
| Any signs of inflammation (discharge, tenderness, fluctuation) | 1 | 5 | 4.067 | 0.35 – 46.65 | 0.30 |
| ***Laboratory Variables at baseline*** | | | | | |
| Baseline Hemoglobin | 12.25 | 12.2 |  |  | 0.874 |
| Baseline ALC | 1460 | 1628 |  |  | 0.294 |
| Baseline ESR | 38.5 | 32 |  |  | 0.683 |
| Baseline CRP | 16 | 11.1 |  |  | 0.325 |
| Baseline CD4 | 719 | 657 |  |  | 0.906 |
| ***Microbiological Variables at baseline*** | | | | | |
| AFB smear positivity at baseline | 1 | 7 | 2.67 | 0.24 – 29.26 | 0.406 |
| Xpert TB Positivity at Baseline | 4 | 29 | ∞ |  | 0.115 |
| Culture Positivity at baseline | 3 | 31 | 3 | 0.29 – 30.44 | 0.614 |
| ***Histopathological Variables at baseline*** | | | | | |
| Well-formed granulomas | 2 | 51 | 0.216 | 0.027 – 1.701 | 0.171 |
| Necrosis | 2 | 58 | 0.069 | 0.008 – 0.626 | 0.039 |

**Table 5: Correlation of Transcription factor T-bet** **with signature cytokine of Th1 immune response (supplementary information)**

Correlation of Transcription factor **T-bet** with signature cytokine of Th1 immune response IFN-γ and TNF-α in the Paradoxical Group (PR-9) vs Non-Paradoxical Group (NPR-13)

| **Timeline** | **Signature cytokine of Th-1 immune response** | **PR** | | **NPR** | |
| --- | --- | --- | --- | --- | --- |
|  | IFN-γ | r | P Value | r | P Value |
| Baseline |  | 0.785 | 0.0121* | 0.600 | 0.030* |
| 2 months Rx |  | 0.22 | 0.5695 | 0.722 | 0.005* |
| 6 months Rx |  | 0.002 | 0.9947 | 0.954 | 0.0001* |
| Baseline | TNF-α | 0.966 | 0.0001* | 0.977 | 0.0001* |
| 2 months Rx |  | 0.863 | 0.0027* | 0.994 | 0.0001* |
| 6 months Rx |  | 0.395 | 0.2916 | 0.954 | 0.0001* |

PR: Paradoxical Group (n-9), NPR: Non-Paradoxical Group (n-13), Correlation coefficient – r, Statistical significance was set as *P* < 0.05*

**Table6: Correlation of Transcription factor GATA-3 with signature cytokine of Th2 immune response (supplementary information)**

Correlation of Transcription factor **GATA-3** with signature cytokine of Th2 immune response **IL-4** and **IL-10** in the Paradoxical Group (PR-9) Vs Non-Paradoxical Group (PR-13)

| **Timeline** | **Signature cytokine of Th-2 immune response** | **PR** | | **NPR** | |
| --- | --- | --- | --- | --- | --- |
|  | IL-4 | r | P Value | r | P |
| Baseline |  | 0.2889 | 0.450 | 0.464 | 0.109 |
| Two months Rx |  | -0.0219 | 0.955 | 0.806 | 0.0009* |
| Six months Rx |  | 0.1822 | 0.638 | 0.8662 | 0.0001* |
| Baseline | IL-10 | 0.4457 | 0.229 | 0.257 | 0.395 |
| Two months Rx |  | 0.6083 | 0.082 | 0.554 | 0.004* |
| Six months Rx |  | 0.1617 | 0.677 | 0.614 | 0.025* |

PR: Paradoxical Group (n-9), NPR: Non-Paradoxical Group (n-13), Correlation coefficient – r, Statistical significance was set as *P* < 0.05*.
